# Triple network model of brain connectivity changes related to adverse mood effects in an oral contraceptive placebo-controlled trial

**DOI:** 10.1101/2022.08.11.22278664

**Authors:** Esmeralda Hidalgo-Lopez, Jonas Engman, Inger Sundström Poromaa, Malin Gingnell, Belinda Pletzer

## Abstract

Combined oral contraceptives (COC) are among the most commonly used contraceptive methods worldwide, and mood side effects are the major reason for discontinuation of treatment. We here investigate the directed connectivity patterns associated with the mood side effects of an androgenic COC in a double-blind randomized, placebo-controlled trial in women with a history of affective COC side effects (n=34). We used spectral dynamic causal modeling on a triple network model consisting of the default mode network (DMN), salience network (SN) and executive central network (ECN). Within this framework, we assessed the treatment-related changes in directed connectivity associated with adverse mood side effects. Overall, during COC use, we found a pattern of enhanced connectivity within the DMN and decreased connectivity within the ECN. The dorsal anterior cingulate cortex mediates an increased recruitment of the DMN by the ECN during treatment. Mood lability was the most prominent COC-induced symptom and also arose as the side effect most consistently related to connectivity changes. Connections that were related to increased mood lability showed increased connectivity during COC treatment, while connections that were related to decreased mood lability showed decreased connectivity during COC treatment. Among these, the connections with the highest effect size could also predict the participants’ treatment group above chance.

**Significance Statement:** Mood side effects are the major reason for discontinuation of oral contraceptive (OC) use. However, the neural substrate underlying these adverse mood effects is still unknown. Here, we investigate the connectivity changes during OC use in women with a history of OC-related mood side effects. We show that mood lability experienced during OC use relates to connectivity patterns previously reported across different mood disorders. The dorsal anterior cingulate cortex, crucial for emotional and cognitive regulation, arose as a mediator area between networks. These neural dynamics during OC treatment may affect cognitive processes underlying mood stability. These findings have important implications for women’s mental health and well-being.

## Introduction

Combined oral contraceptives (COC) are among the most commonly used contraceptive methods worldwide (POP/DB/CP/REV 2019). First introduced in the 1960’s, nowadays more than 151 million women of reproductive age use this method, and more than 80% of women from the US have used it at some point in their life (1). COCs contain a synthetic estrogen, commonly ethinyl-estradiol, and a progestin that can be classified as androgenic or anti-androgenic, depending on its interaction with the androgen receptor (2, 3). These exogenous hormones exert, via negative feedback, downregulation of the hypothalamic–pituitary–gonadal axis, and therefore, decrease the endogenous ovarian hormone production.

Although research is still scarce, the continued exposure to synthetic progestins, has been proposed to underlie the adverse mood effects that some women experience during COC treatment (4, 5). While observational studies come to different conclusions as to the risk of developing affective disorders (5, 6), mood side effects are the major reason for discontinuation of COC treatment (7, 8). Randomized, placebo-controlled trials (RCTs) on adverse mood symptoms are rare, but the prevalence of this side effect is estimated to be between 4-10%, particularly for androgenic COC (8). In a well powered RCT, Zethraeus et al. (9) tested the effect of an androgenic COC (levonorgestrel), which decreased vitality, well-being and self-control compared to placebo. Lundin et al., (4) carried out an RCT with a partially anti-androgenic COC (nomegestrolacetate), with premenstrual improvement in depression but similar adverse inter-menstrual mood-related effects, such as enhanced irritability and mood lability compared to placebo. At the same time, anti-androgenic COCs may have beneficial effects on mood, especially for women with pre-menstrual symptoms (10). Thus, the susceptibility for adverse mood effects during COC-treatment varies between subjects. However, up to now little is known about individual risk factors.

Although mood-related side effects are usually more present in women with a history of depressive symptoms, depressed mood is not consistently reported to change after treatment in RCTs (9, 11). These depressive symptoms may, however, appear in the long term, given that prospective cohort studies have associated the use of COC (and hormonal contraception in general) to the subsequent use of antidepressants and depression diagnosis (5). Impaired emotion discrimination, reactivity and response have been suggested to underlie the adverse mood-related effects of COC (see review 12). For instance, COC users show deficient fear extinction (13), impaired emotion recognition (14, 15) or differential emotional memory (16, 17), all processes related to the symptomatology of mood disorders.

Recent years have witnessed an increased integration of psychiatry and neurosciences. This has allowed us to change our perspective on mental illnesses and further understand the underlying neural correlates, which, eventually, improves prevention, diagnosis and treatment of disorders. Within this approach, the triple network model, seeks to relate major psychopathologies to differences in large-scale brain organization (18). Intrinsic brain networks are characterized by temporally correlated activity at rest, which can be identified by functional magnetic resonance imaging (fMRI). According to the triple network model, a balanced connectivity between three ‘core’ networks, i.e. the default mode network (DMN), the executive control network (EN) and the salience network (SN), is crucial for mental health. These three networks support basic and higher-order regulatory functions fundamental to information processing and are thus essential to a variety of cognitive and emotional functions. The DMN is related to self-referential mental activity and increases its activity during the resting state. Its major nodes include the precuneus/posterior cingulate cortex (PCC), bilateral angular gyrus (AG) and medial prefrontal cortex (mPFC) (19). The ECN is a frontoparietal network related to the active manipulation and inhibition of information and thus associated with planning, decision making, and the control of attention and working memory (20). It includes the bilateral middle frontal gyri (MFG) and bilateral supramarginal gyri (SMG). The SN is important for detecting and integrating salient external and internal stimuli (20). It is comprised by bilateral anterior insula (AI) and dorsal anterior cingulate cortex (dACC). The SN, especially the insular cortex, acts as a switcher between networks and has been characterized in healthy population as engaging the ECN in response to relevant stimuli (21).

Deficits in dis/engagement between the nodes of these networks are suggested to be particularly relevant for affective disorders (18). With regards to emotional stimuli, this could be a weakened salience map and aberrant filtering of stimuli (18), and both depression, social anxiety and panic disorder have been associated with reduced connectivity within the SN, together with hyper-connectivity between the DMN (especially medial areas) and the SN (22). For the ECN, hypo-activation has been found in frontal areas of both depressed and anxious patients, while hyper-connectivity between frontal nodes of the ECN and SN (MFG-ACC) has been suggested to reflect a compensatory mechanism (22). Neuroimaging studies have found differences between naturally cycling women and COC users in structure and function of areas belonging to these three networks and related these differences to emotional and cognitive processing (for a review, see 23). Therefore, we propose the triple network model as a valuable framework to study the COC-dependent modulation of connectivity patterns particularly in women experiencing adverse mood side effects.

We here aim to disentangle the directed connectivity patterns associated with the mood side effects of an androgenic COC. To address this question, we used spectral dynamic causal modeling to analyse data from a double-blind RCT, for which women were selected based on previous adverse mood effects on COC and where the COC group experienced increased mood lability, fatigue and depressed mood compared to the placebo group (11). Given that previous research hints at an enhanced connectivity within the DMN (especially medial areas), but decreased connectivity within the ECN related to depressive symptoms (22, 24), we expect this pattern of within-network connectivity also during COC treatment in comparison to placebo, and that the changes in the pattern should be related to depressive mood. Regarding between-network connectivity, functional connectivity findings from this dataset already show an increased connectivity between the medial nodes of the SN and DMN (dACC and PCC) during COC treatment (25), though the directionality of this effect remains unclear. Using DCM among the nodes of the triple network model, will allow us to resolve the directionality of this interaction and other interactions between the SN and DMN, and also explore the directed connectivity of these nodes to the ECN. Related to a potential impaired filtering of stimuli, we also expect a weaker engagement of the frontal ECN by the SN (18) and decreased afferent connectivity into the SN during COC treatment (26, 27).

## Results

In general, in the COC group compared to the placebo group, the within-network connectivity increased during treatment in the DMN, whereas it decreased in the SN and ECN.

Regarding the between-network connectivity, specifically from the dACC (SN) to medial nodes of DMN (PCC and mPFC) effective connectivity was increased in the COC group compared to the placebo group. From the rAG (posterior DMN) to the SN, effective connectivity increased in the placebo group compared to the COC group. Conversely, effective connectivity increased from the rAG to the posterior ECN in the COC group compared to the placebo group.

Effective connectivity from the frontal ECN to the DMN was in general stronger during treatment in the placebo than in the COC group, while those connections originating in the lSMG followed the opposite pattern. Effective connectivity between the ECN and the SN was stronger in the COC group compared to the placebo group.

Given that the placebo group connectivity was also subject to change due to the endogenous hormonal fluctuations of the natural menstrual cycle, we considered whether these interactive effects where due to changes a) only in the COC group, b) in opposite direction for the COC and the placebo group or c) only in the placebo group. Results according to this classification are illustrated in Fig. 2 and summarized in Table 1. When making this distinction some consistent patterns could be distinguished in the relation to the side effects (see Table 1).

**Table 1.** Summary of connections that showed interactive effect of session*treatment.

|  | Session*<br>treatment | Group |  | Side effects<sup>1</sup> |  |  |
| --- | --- | --- | --- | --- | --- | --- |
|  |  | COC | Placebo | Mood lability | Fatigue | Depressed mood |
| From PCC to right AG | + | + |  | + | + |  |
| From left AI to right AG | + | + |  | - |  |  |
| From dACC to left SMG | + | + |  | + | + | - |
| From right AG to PCC | + | + |  |  | - | + |
| From right AG to left SMG | + | + |  | + | - |  |
| From right AG to right SMG | + | + |  | + | - |  |
| From left AG to right MFG | + | + |  | + | + |  |
| From left AG to right AI | + | + |  | - |  | + |
| From left AG to mPFC | + | + | - | + |  |  |
| **From dACC to PCC** | **+** | **+** | **-** | **+** |  | **-** |
| From dACC to mPFC | + | + | - | + | - |  |
| From left SMG to PCC | + | + | - | - |  |  |
| **From right MFG to dACC** | **+** | **+** | **-** | **+** |  | **+** |
| **From right AG to dACC** | **-** | **-** |  | **-** | **-** |  |
| From right SMG to left MFG | - | - |  | - |  | + |
| From left MFG to right AG | - | - |  | - |  | + |
| From left MFG to PCC | - | - |  | - | - | + |
| From right MFG to right SMG | - | - | + | + |  |  |
| From right AI to mPFC | - | - | + | - | + | - |
| dACC to itself | - | - | + | - |  |  |
| From right AG to mPFC | - |  | + |  | - | + |
| From right AG to left AI | - |  | + | - | - | - |
| **From right AG to right AI** | **-** |  | **+** | **-** | **-** | **-** |
| From left AI to right AI | - |  | + |  | + |  |
| From right AI to PCC | - |  | + | - |  | - |
| From left MFG to left AG | - |  | + | - |  | + |
| From left MFG to mPFC | - |  | + | - | - | - |
| From right MFG to left AG | - |  | + | - | - |  |
| From right MFG to right AG | - |  | + |  | + | - |
| From right SMG to right MFG | - |  | + |  |  |  |
| From left SMG to left AG | + |  | - | + |  |  |
| From left SMG to right AG | + |  | - | - | - | + |
| From right SMG to left AI | + |  | - |  | - |  |
| From right SMG to right AI | + |  | - |  |  | - |
| From left MFG to left SMG | + |  | - |  |  |  |
| From PCC to rAI | + | - | - - | + |  |  |

All connections survived a 95% posterior probability threshold and an estimated value of 0.10.
Columns 3-4 correspond to connectivity changes present during COC or placebo treatment respectively, indicated by a + if the connectivity increased and - if the connectivity decreased. Side effects modulation of the treatment connectivity are detailed in columns 5-7, indicated by a + if relationship to the connectivity parameter was positive, and - if the relationship was negative. Connections that were able to predict individual treatment are marked in bold.
<sup>1</sup> Relationship between post-treatment connectivity and scaled difference between the last week of the treatment compared to the correspondent pre-treatment week.

**Figure 1.**
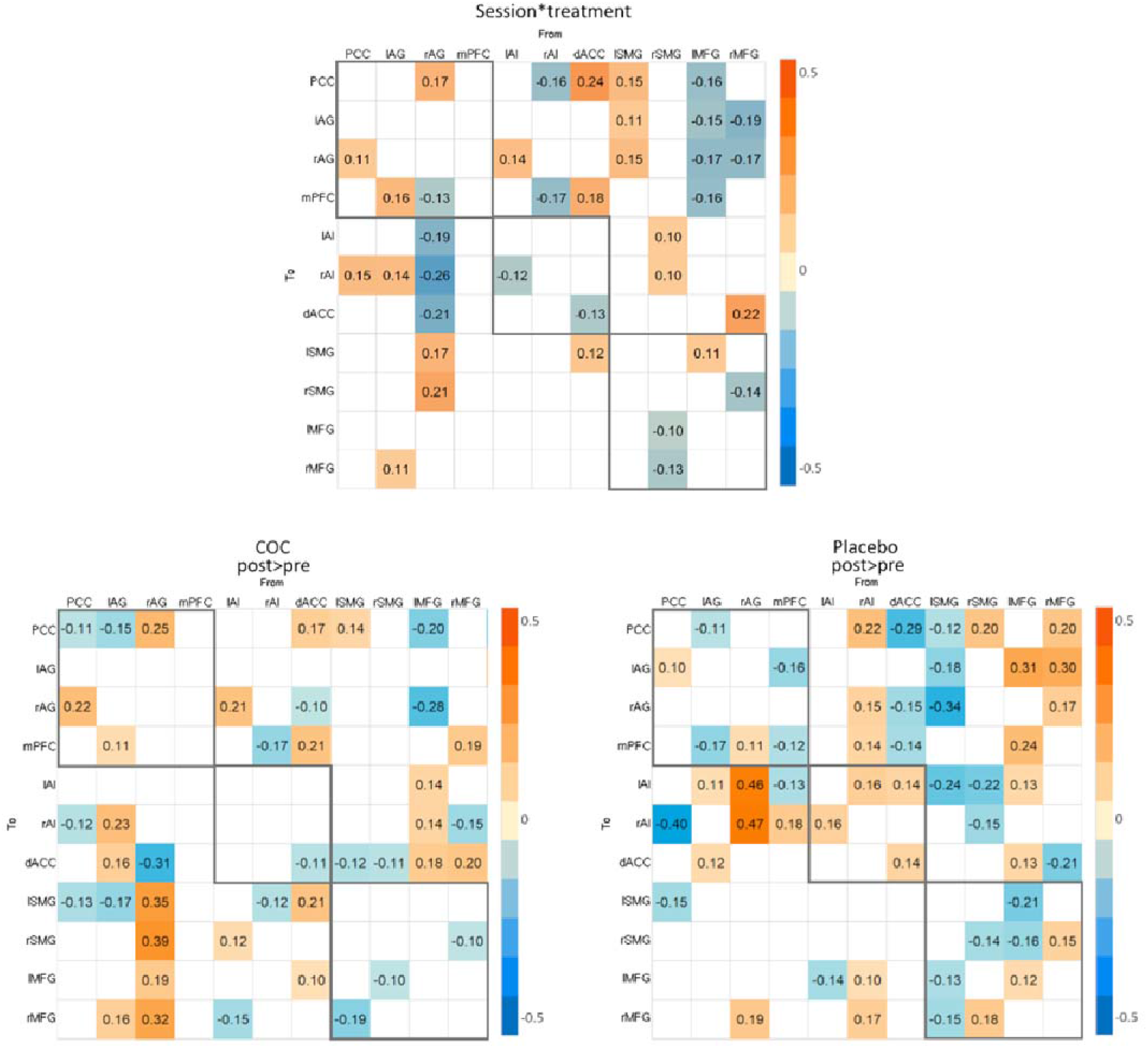
Estimated parameters for a) the interactive effect of session*treatment, b) COC treatment effect, c) placebo effect. These connections surpassed a posterior probability of 95% and an estimated value (Ep) of 0.10. The exact Ep is indicated in each cell, warm colours indicating positive parameter estimates and cold colours negative. The columns are the outgoing connections, and the rows are the incoming connections, ordered as: PCC, lAG, rAG, mPFC, lAI, rAI, dACC, lSMG, rSMG, lMFG, and rMFG

**Figure 2.**
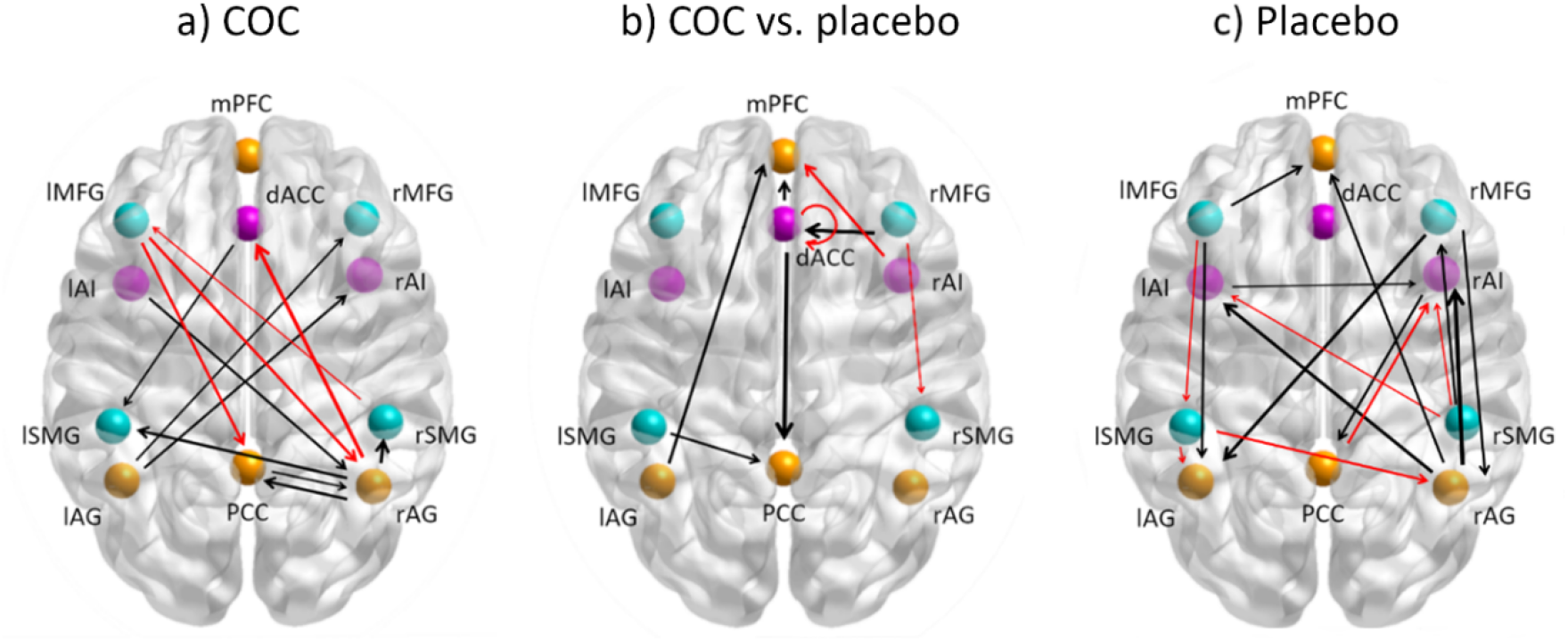
Interactive effects of session*treatment due to changes a) only in the COC group, b) in opposite direction for the COC and the placebo group or c) only in the placebo. These connections surpassed a posterior probability of 95% and an estimated value of 0.10. The differential connectivity strengths (Ep) are depicted by the width of the arrow. Black arrows reflect positive values and red arrows reflect negative values.

### Associations of effective connectivity mood side effects

Connections increasing in the COC group, independently of whether they also decreased in the placebo group or not, were in general positively related to mood lability. Connections decreasing in the COC group, independently of whether they also increased in the placebo group or not, were in general negatively related to mood lability. For those connections increasing only in the placebo group, we found mostly negative relationship to mood lability, while for those connections decreasing only in the placebo group, no consistent pattern of association to mood side effects was found (Table 1, Fig. 3).

**Figure 3.**
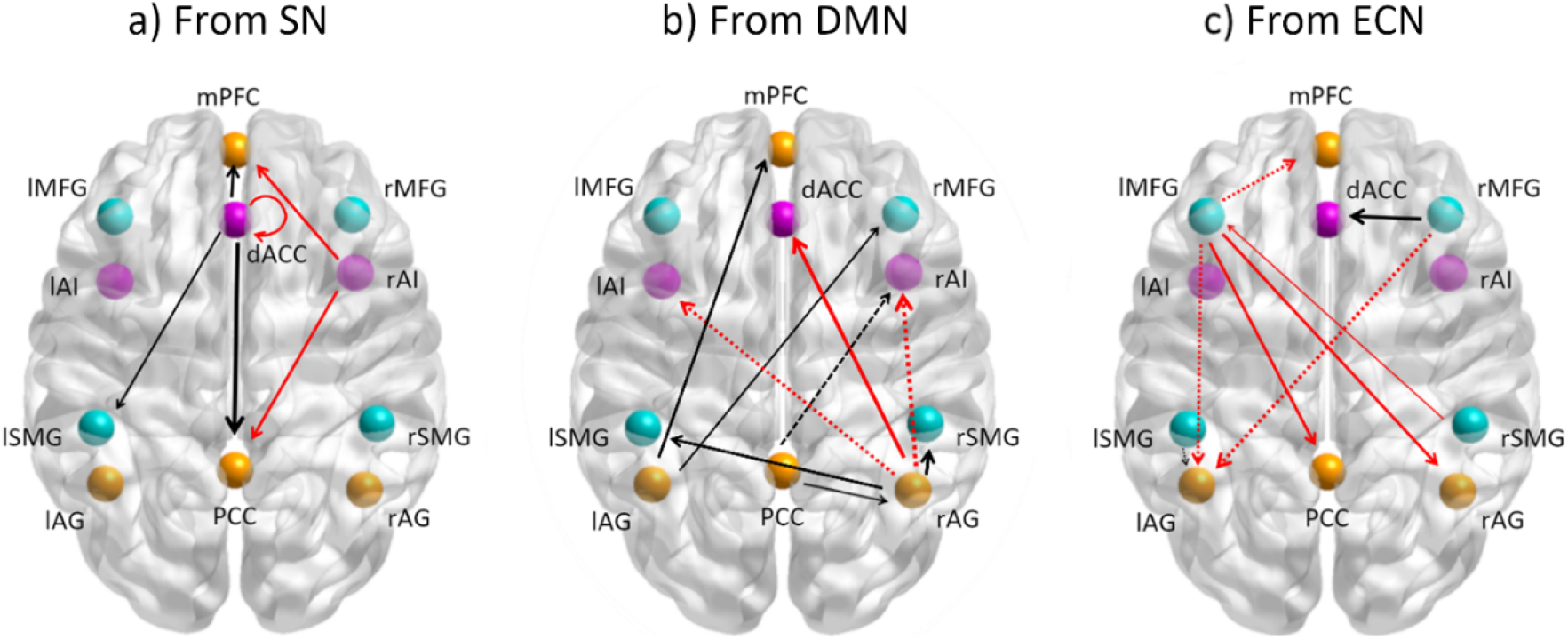
Connections showing relationship to mood lability differences a) from the SN, b) from DMN c) from ECN. These connections surpassed a posterior probability of 95% and an estimated value of 0.10 and related during treatment to the mood-lability changes from the previous cycle, in the same direction as they changed for the COC-treatment. The differential connectivity strengths (Ep) are depicted by the width of the arrow. Black arrows reflect positive values and red arrows reflect negative values. Dashed connections were those that only changed in the placebo group from follicular to luteal phase.

Among those mood lability-related connections, the following connectivity changes surpassed the threshold of Ep>0.20: from dACC to PCC, from rMFG to dACC, from rAG to rAI, from rAG to dACC, and from rAG to rSMG. In order to maximize the predictive accuracy, only these connections were selected in the subsequent cross-validation analysis.

### Prediction of treatment by mood-related effective connectivity

The leave-one out cross-validation based on those connections identified above as showing the highest effect size and relation to mood lability (dACC → PCC, rMFG → dACC, rAG → rAI, rAG → dACC, rAG → rSMG) showed a significant association between the actual and predicted group of *r*_df:66_=0.22, *p*=0.03 (Fig. 4).

**Figure 4.**
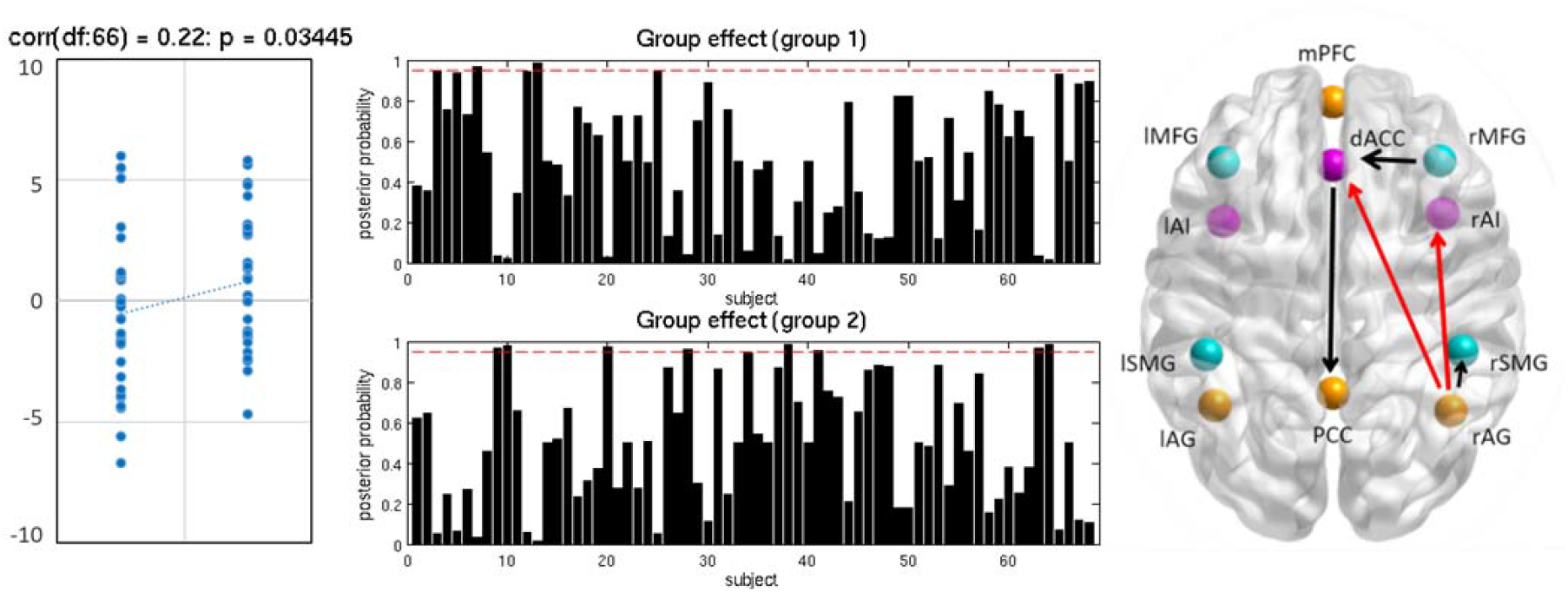
Leave-one-out cross-validation analysis. Left: scatter plot displaying the correlation between the actual treatment group in the left-out-subject’s design matrix and the predicted treatment group based on the left-out-subject’s connectivity. Centre: the resulting posterior probability for each treatment group for each subject. Right: Differential connectivity strength for the interactive effect of session*treatment. The differential connectivity strengths (Ep) are depicted by the width of the arrow. Black arrows reflect positive values and red arrows reflect negative values.

## Discussion

The main goal of the current manuscript was to characterize the changes in directed connectivity during COC treatment related to concurrent mood symptoms. Our results showed how effective connectivity changes noted during COC treatment related to mood deterioration. Mood lability was the most prominent COC-induced symptom (11), and also arose as the side effect most consistently related to connectivity changes. Connections that were related to increased mood lability showed increased connectivity during COC treatment, while connections that were related to decreased mood lability showed decreased connectivity during COC treatment. Among these, the connections with the highest effect size could also predict the participants’ treatment group above chance.

Overall, we found the expected pattern of enhanced connectivity within the DMN and decreased connectivity within the ECN during COC use (22, 28). Increased connectivity within the DMN has been related to ruminative thoughts and lower connectivity within the ECN to lower cognitive control (see reviews,28, 29). Specifically for the posterior DMN, increased within-network connectivity has been observed in major depression patients (30), suggested to underlie ruminative thoughts in this clinical population (29), and been related to premenstrual-like symptoms in COC users (31). Interestingly, after antidepressant treatment, decreased functional connectivity within the posterior sub-network of the DMN, comparable to healthy subjects, has been reported (30). However, and contrary to our hypothesis, only the connectivity from rAG to PCC was positively related to depressive symptoms. In that respect, different sub-items in depressive scales have been shown to be associated with abnormal activity of various brain areas (32). Additionally, depressive symptoms derived from chronic versus short time COC use may differ. Clinical major depression usually has a rather slow onset, and it is highly likely that our treatment period of three weeks is too short to fully assess the development of depressive symptoms.

Alongside the decreased ECN within-network connectivity, which is line with previous findings (33), we observed a clear distinction in ECN-DMN between-network connectivity dynamics along the anterior-posterior axis. While connectivity from the left frontal ECN to the DMN decreased in COC-users, there was a bidirectional increase in connectivity between the posterior DMN (pDMN) and parietal ECN (pECN). The frontal nodes of the ECN are key regions for emotional processing and regulation (34, 35), whereas the parietal areas of the ECN and DMN appear to be crucial for the cognitive reappraisal of negative stimuli (see 36 for a meta-analysis). Disrupted activity of the frontal ECN has been observed for depression, anxiety, and bipolar disorder(22, 37–39).

Engagement of the parietal areas in mood and anxiety disorders has been proposed as a compensatory mechanism in response to impaired function of frontal areas (36).Moreover, the frontal ECN has been suggested as a neural marker of clinical responsivity to treatment (40) and has been used as a target during antidepressant treatment with transcranial magnetic stimulation (41). In the present sample, these connectivity changes were in general associated with mood lability but not consistently with depressive symptoms.

Remarkably, an important role seems to emerge for the dACC. Better emotional regulation and lower anxiety levels have been related to stronger dACC activity (42, 43), and depressive symptoms are associated with reduced dACC volume (44). More importantly, this area has been identified as neural predictor of individual therapy response in depression (40) and anxiety disorders (45, 46). In the present sample, the dACC seems to act as a mediator between the DMN and ECN. During COC-treatment, the medial nodes of the DMN are recruited by the dACC (compare also 25), while the dACC in turn is recruited by the frontal ECN. The over-recruitment of the ACC by the medial DMN has been described in major depressive (28) and anxiety disorders (22) in relation to impaired attention to relevant stimuli (18). In healthy adults resting-state functional connectivity between PCC and ACC was positively related to both negative and positive daily mood and interpreted as a general enhanced reactivity to arousal as opposed to valence (47). Regarding the engagement of the dACC to the frontal ECN,, MFG-ACC hyper-connectivity has been suggested to reflect an increase in top-down control (48) as a compensatory mechanism in anxiety and depressive disorders (22). It is worth noting that the pivotal role of the dACC seems also important for the group treatment prediction in the present sample.

The above-described changes and related experienced side effects could be a consequence of the synthetic hormones, the abolishment of cyclic endogenous hormonal fluctuations, or both. Although animal research shows a differential binding affinity of synthetic compared to endogenous hormones (levonorgestrel has a 5-fold affinity vs. progesterone for progesterone receptors) (2), the main neuroactive effects of progesterone are mediated by its metabolites and its interaction with GABA-receptors. Relatedly, long term contraceptive treatment in rodents (4-6 weeks including levonorgestrel) has shown to reduce cerebral progesterone and allopregnanolone and increase anxiety-like behaviour, related to changes in GABA-receptor (49, 50). Moreover, low endogenous estradiol levels during COC use have been previously related to impaired fear extinction in a cross-species experiment with female rats and women (13), while high estradiol levels during the peri-ovulatory phase in naturally cycling women have been related to an advantage for emotion processing (51). Therefore, we find it likely that the constant enhanced synthetic progestins effects, alongside the absence of endogenous hormonal cyclicity and estradiol-progesterone interaction may be of high relevance for the mood side effects and related changes in connectivity here described.

Although it is out of the scope of this study, connectivity changes experienced by the placebo group along their menstrual cycle in this study did not completely match changes in healthy naturally cycling women reported in our previous work (52). Although the main reason for these discrepancies may be differences in the time window of each appointment, individual variability in functional responsivity to sex hormones could have also contributed (53). While the present study deliberately included women who had previously reacted with mood-effects to COC, they were also allowed to report experiencing premenstrual syndrome (PMS), while this condition was an exclusion criterion in the previous study. Accordingly, some of these differences could relate to both the vulnerability in women for PMS and the susceptibility to adverse mood effects during COC use.

In summary, the present randomized placebo-controlled trial showed effective connectivity changes during COC treatment related to worsened mood in women with a history of mood COC side effects. The most confident effects corresponded to connections that changed during COC treatment compared to placebo and were related to an increased in mood lability. These differences during COC treatment in the triple network model may affect cognitive processes important for mood stability and mental health and similar disruptions have been reported across mood disorders (18, 22). Further studies are needed in order to shed light on specific mechanisms by which synthetic hormones exert changes on these neural substrates on one hand, and which specific features correspond to an increased vulnerability to experience adverse mood side effects on the other. The paucity of research regarding the neuroactive effects of COC calls for more extensive research, especially when considering the time window when women start using COC, due to the plasticity in adolescent brains.

## Materials and Methods

### Participants

Thirty-four healthy women with previously reported COC-induced mood deterioration participated in a double-blinded, randomized, parallel-group clinical trial. During one treatment cycle, the participants received either a COC (ethinyl estradiol (EE) 30 mg/0.15 mg levonorgestrel, provided by Bayer Pharma AB) or placebo (Bayer Pharma AB). Results from task-based paradigms and salience resting-state functional connectivity have been reported and discussed elsewhere, with additional details about the study design and participants demographics (11, 25, 54). Exclusion criteria were the use of hormonal contraceptives, cortisol, levothyroxine, or psychotropic drugs within the previous two months; and neurological or psychiatric disorders. All methods conform to the Code of Ethics by the World Medical Association (Declaration of Helsinki) and were approved by the Independent Research Ethics Committee, Uppsala University and the Medical Products Agency in Sweden. EU Clinical Trial Register (https://www.clinicaltrialsregister.eu), number: 2008-003123-24.

### Experimental design

Participants were scanned twice, first during a pre-treatment cycle (day 1—10 after onset of menses) and secondly, during the last week of the treatment cycle (day 15—21 after start of treatment). The participants started taking the pill on the first day of menses (Fig 5). In the COC group serum concentrations of ethinyl estradiol and levonorgestrel were expected to be stable during treatment, with low levels of endogenous hormones. However, at the second scan, the placebo group was expected to have an increase in endogenous hormone levels (which, for most individuals, coincided with the luteal phase). Blood samples were drawn at each of the scanning sessions for hormonal analyses. After centrifugation and storage at -70 °C, samples were analyzed using a Roche Cobas e601 and Cobas Elecsys reagent kits (Roche Diagnostics, Bromma, Sweden). Additionally, participants filled out daily mood and physical symptoms on the Cyclicity Diagnoser (CD) scale (55) both during the pre-treatment and treatment cycles. In order to control for the effect of possible pre-menstrual syndrome (PMS) symptoms on mood-ratings, changes pre-and during treatment were assessed as the standardized difference between the last week of the treatment compared to the corresponding pre-treatment week (i.e. in both cases during the luteal phase).

**Figure 5.**
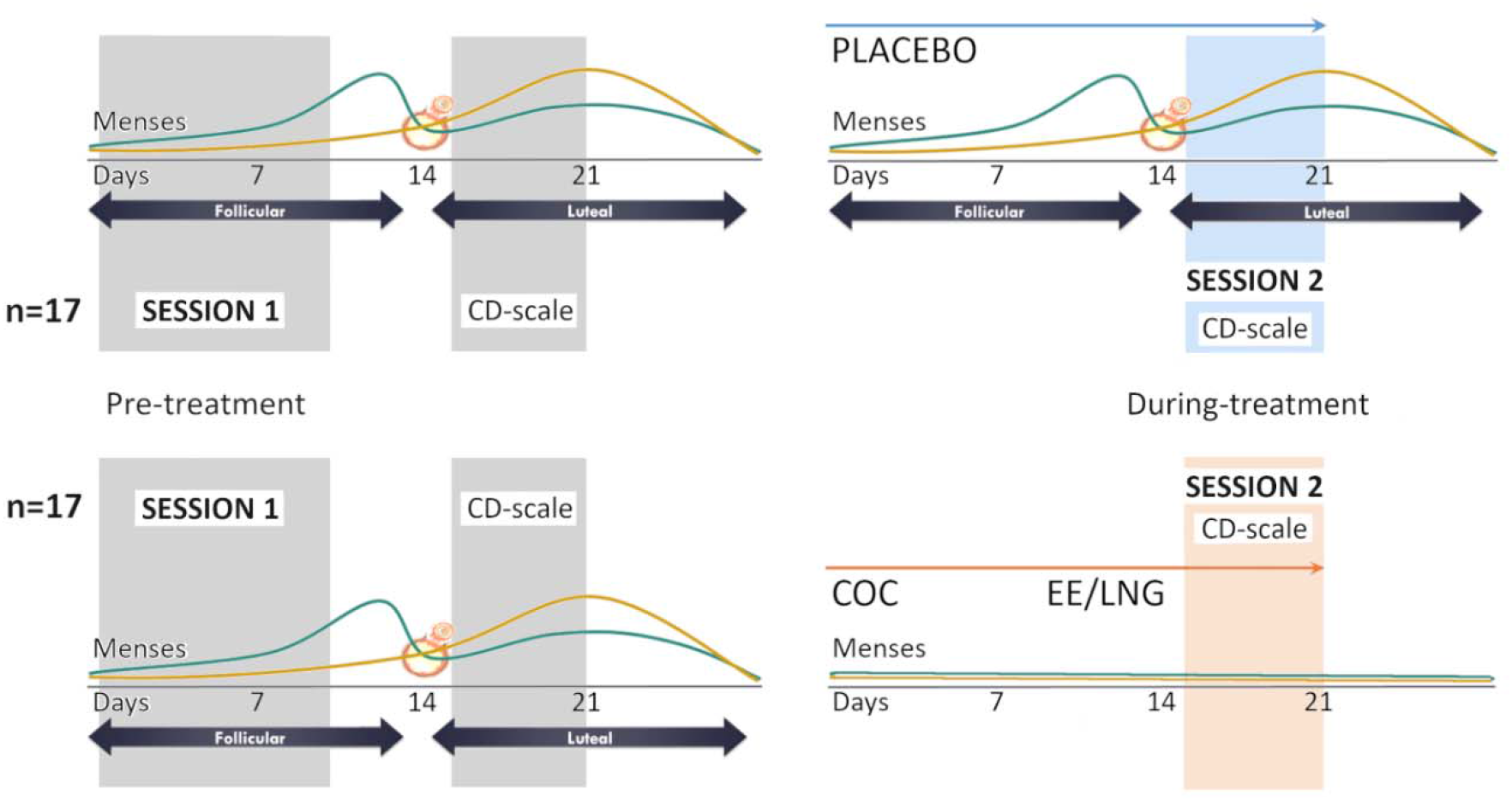
Experimental design: Each participant had two sessions, before and during treatment (day 1—10 and day 15—21 respectively after onset of menses). Therefore, for the placebo group endogenous hormone levels increased from the first to the second appointment, while in the COC group synthetic hormone levels were stable, and endogenous hormones low.

### Data acquisition

Functional and structural images were acquired on a Philips Achieva 3.0T scanner using an 8-channel head coil (Philips Medical Systems, Best, The Netherlands). For the 5 min resting state a single shot echo planar imaging sequence was used to collect 100 volumes of BOLD data with a voxel size of 3.0×3.0×3.0 mm^3^ in 30 ascending slices (TR=3000ms, TE=35ms, flip angle=90°, and FOV= 230×230 mm^2^). Participants were instructed to stay as still as possible and simply rest with their eyes closed. For the structural images an inversion recovery turbo spin echo sequence was used to acquire a structural T1-weighted image with a voxel size of 0.8× 1.0×2.0mm^3^ in 60 slices (TR=5700ms, TI=400ms, TE=15ms, and FOV=230×230mm^2^).

### Preprocessing

Scanner DICOM images were first converted to nifti files with MRIcron (www.nitrc.org/projects/mricron/). Images were pre-processed using SPM12 standard procedures and templates SPM12 (www.fil.ion.ucl.ac.uk/spm) and despiked using 3D-despiking as implemented in AFNI (afni.nimh.nih.gov). Pre-processing included realignment of the functional images, segmentation of the structural images, co-registration of the functional images to the structural images, normalization of functional images using the normalization parameters and spatial smoothing using a 6 mm kernel. The resulting images were subjected to the ICA-AROMA algorithm implemented in fsl including non-aggressive removal of artifactual components (56).

### Selection and extraction of volumes of interest

Eleven ROIs were selected as core nodes of the corresponding networks, based on a large body of literature. First, for the DMN, the precuneus/posterior cingulate cortex (PCC), bilateral angular gyri (AG) and medial prefrontal cortex (mPFC) (57, 58). Second, for the SN, bilateral anterior insula (AI) and dorsal anterior cingulate cortex (dACC) (20, 58). Third, for the ECN, bilateral middle frontal gyri (MFG) and supramarginal gyri (SMG) (59). ROI specific masks were created with the Wake Forest University (WFU) Pickatlas toolbox (60). Using spatial ICA as implemented in the Group ICA for fMRI Toolbox (GIFT, http://mialab.mrn.org/software/gift), group-level peaks for each ROI were identified within each intrinsic connectivity network (ICN) (61). For this, we first extracted 20 components, identified each of them via spatial correlation to pre-existing templates (62), and selected those four corresponding to the DMN, SN, left ECN and right ECN. Functional connectivity between the ROIs was further corroborated to be positive between ROIs of the same network and anti-correlated between different networks using the CONN toolbox (63). The ROIs, their functional connectivity and their group-level peak coordinates are listed in Table 2 and shown in Figure 6. In order to extract the principal eigenvariate from each of the 11 ROIs, subject-specific coordinates were determined as local maximum within 8 mm of the group-level coordinates, but restricted to still be within the ROI specific mask. The time series from each ROI were then used in subsequent DCM analyses.

**Table 2:**
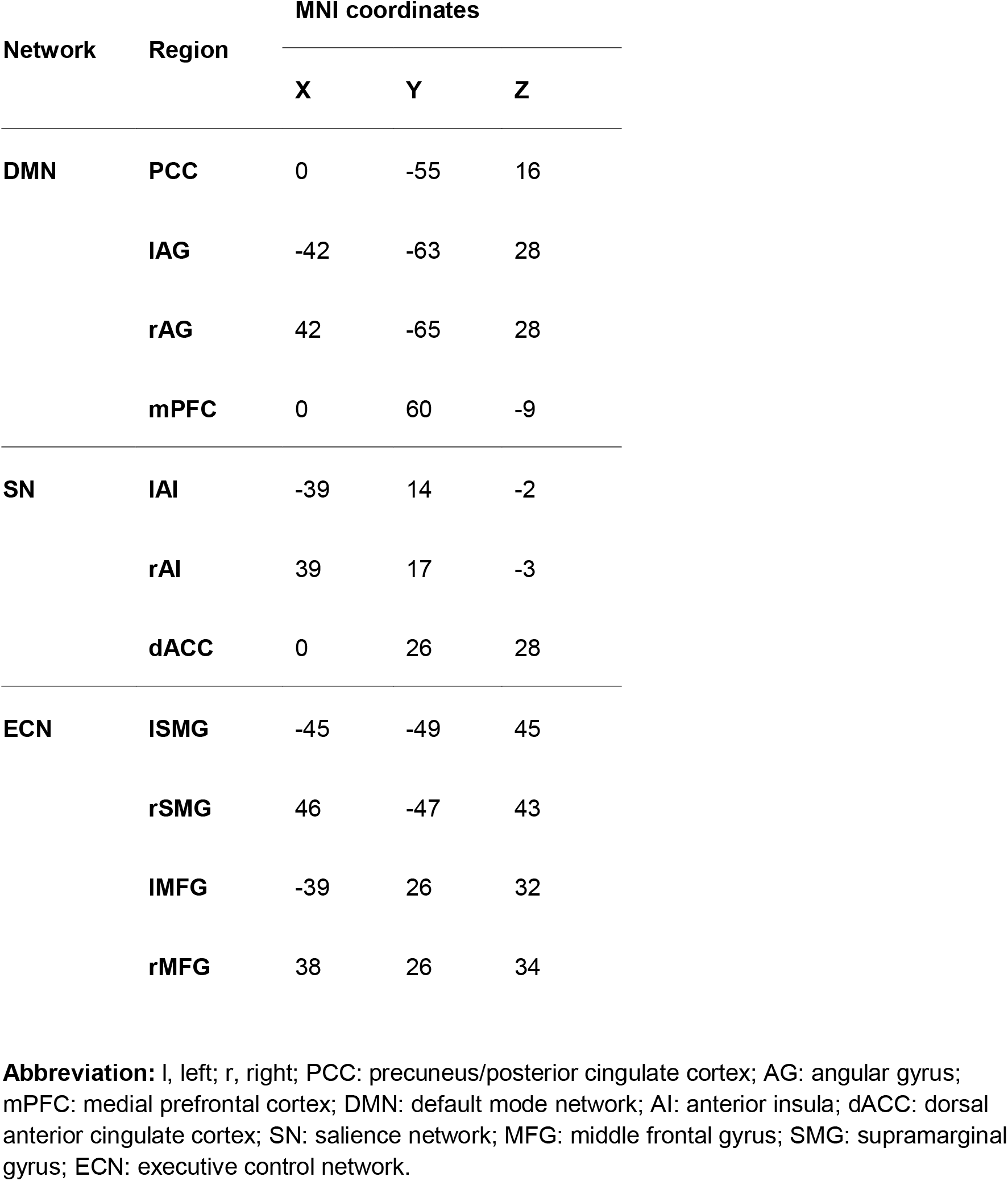
Group level coordinates for each region of interest.

**Figure 6.**
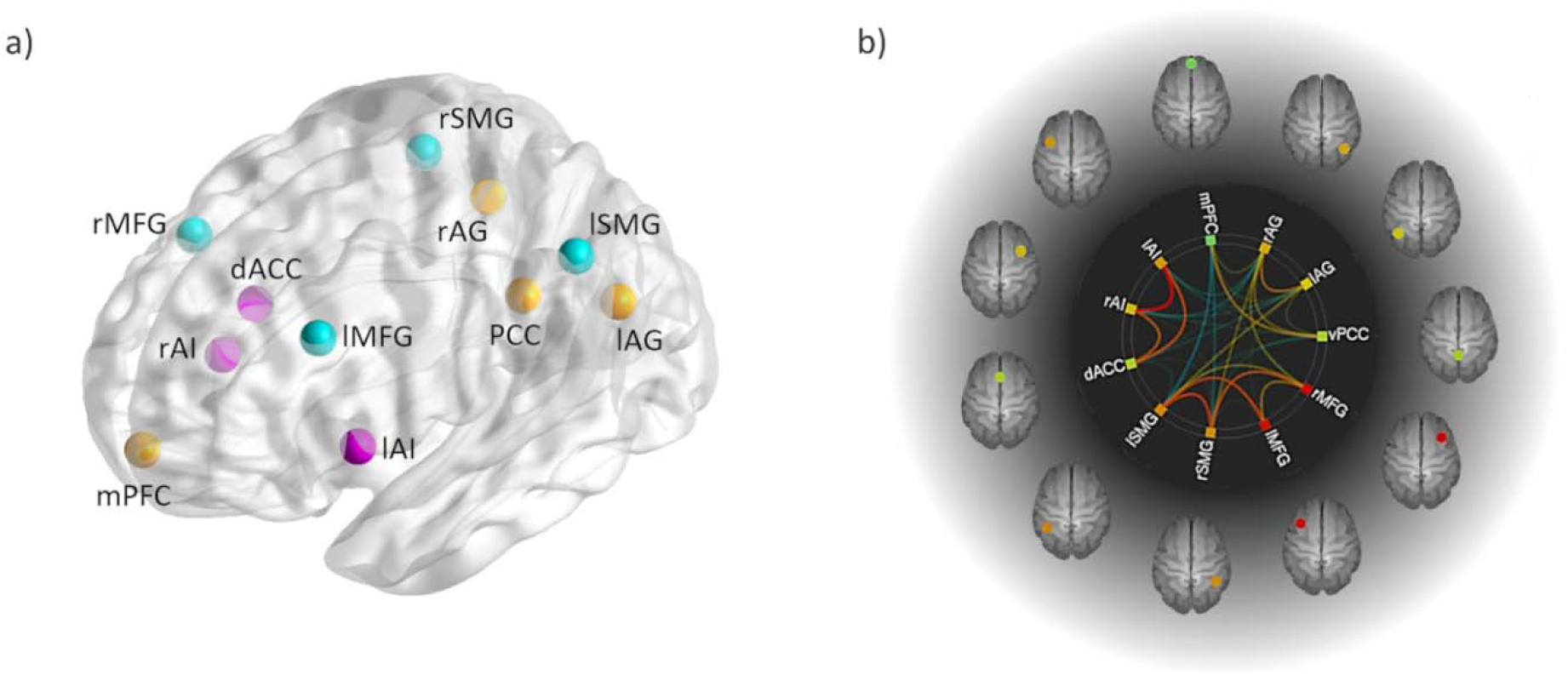
Regions of interest for each network and functional connectivity: The regions of interest (ROIs) used in the current study is shown in (a), corresponding to default mode brain regions (yellow): precuneus/posterior cingulate cortex (PCC), medial prefrontal cortex (mPFC) and bilateral angular gyrus (AG); the salience network (pink): bilateral anterior insula (AI) and dorsal anterior cingulate cortex (dACC); and the executive control network (blue): bilateral middle frontal gyri (MFG) and bilateral supramarginal gyri (SMG). (b) Functional connectivity between the ROIs was positive within the same network (warm colours) and anti-correlated between different networks (cold colours).

### Spectral Dynamic Causal Modeling and Parametric Empirical Bayes

Resting state functional images were modelled using a (Bayesian) hierarchical random effects framework and spectral DCM was specified and inverted using DCM12 as implemented in SPM12 (www.fil.ion.ucl.ac.uk/spm). At the first level, for each participant and session, a fully connected model of 121 parameters (including all possible connections between nodes) was specified to fit the complex cross-spectral density estimating the intrinsic effective connectivity (i.e., the ‘A-matrix’) within and between networks. This estimation takes into account the effects of neurovascular fluctuations as well as noise (64). Default priors implemented in SPM were used at this level and quality of the process was ensured by a variance explained by the model higher than 90% (65).

In order to compare changes in the treatment group to the changes observed in the placebo group, we ran a 2-level hierarchical analysis using a Parametric Empirical Bayes (PEB)-of-PEBs approach. We first modelled the treatment effect on each group separately, and then fit those parameters to the next level of the hierarchy as implemented in the Parametric Empirical Bayes framework in the SPM software (65, 66). In this way, we captured the overall mean connectivity, the main effects of treatment and group, and most relevant, the interaction between group and treatment. This hierarchical approach, with a general linear model (GLM) to capture effects of interest in the second and third level, allowed us to captured more accurately the between subjects’ variability (random effects).

In order to relate during treatment effective connectivity to mood related symptoms, we modelled another PEB including the changes in those CD-scale ratings that differed significantly between the COC and placebo group, i.e. *mood lability, fatigue* and *depressed mood*^1^ (11) (Fig.S1. suppl. material).

PEB results were thresholded to only include parameters from the A matrix that had a 95% posterior probability of being present vs. absent, which represents strong evidence for treatment-related changes, and thresholded to an estimated value Ep>0.10. Only results surviving this threshold are reported in the results section.

### Cross-validation

In order to check whether the mood side effect related effective connectivity could predict the assignment of participants to one group or another (COC vs. placebo) we used a leave-one-out scheme (spm_dcm_loo.m) as described in (67). This way we can assess the association between the actual group in the left-out-subject’s design matrix (pre-post COC-placebo) and the predicted group correspondence based on the left-out-subject’s connectivity. Given that there is a strong dilution-of-evidence effect, we selected those parameters that maximize the predictive accuracy based on their i) effect size and ii) relation to mood lability. Accordingly, we first thresholded the session by treatment interaction to Ep>0.20 and then included only those parameters that were positively related to mood lability when increased during COC treatment and negatively related to this side effect when decreased during COC treatment.

## Data Availability

All data produced in the present study are available upon reasonable request to the authors.

## Acknowledgments

This research was funded by the Swedish Research Council project K2008-54X-200642-01-3, the Swedish Council for Working Life and Social Research projects 2007-1955, and 2007-2116, the Family Planning Foundation, and an unrestricted research grant from Bayer AB. The European Research Council (ERC) Starting Grant 850953 supported BP and EH-L.

## Supplementary material

**Fig.S1. Estimated parameters for mood related symptoms**. These connections surpassed a posterior probability of 95% and an estimated value (Ep) of 0.10. The exact Ep is indicated in each cell, warm colours indicating positive parameter estimates and cold colours negative. The columns are the outgoing connections, and the rows are the incoming connections, ordered as: PCC, lAG, rAG, mPFC, lAI, rAI, ACC, lSMG, rSMG, lMFG, and rMFG.

## Footnotes

1 Please note that the pre-treatment CD ratings were obtained during the luteal phase, while the pre-treatment MRI was performed during menses. This time-point was chosen to avoid potential menstrual-cycle related variations in the control group, while making sure that the changes in the COC group capture only those increases in mood lability, fatigue and depressed mood that surpass potential menstrual cycle related variations.

