## Supplemental Fig S1 for "Triple network model of brain connectivity changes related to adverse mood effects in an oral contraceptive placebo-controlled trial"

**SUPPLEMENTARY MATERIAL**

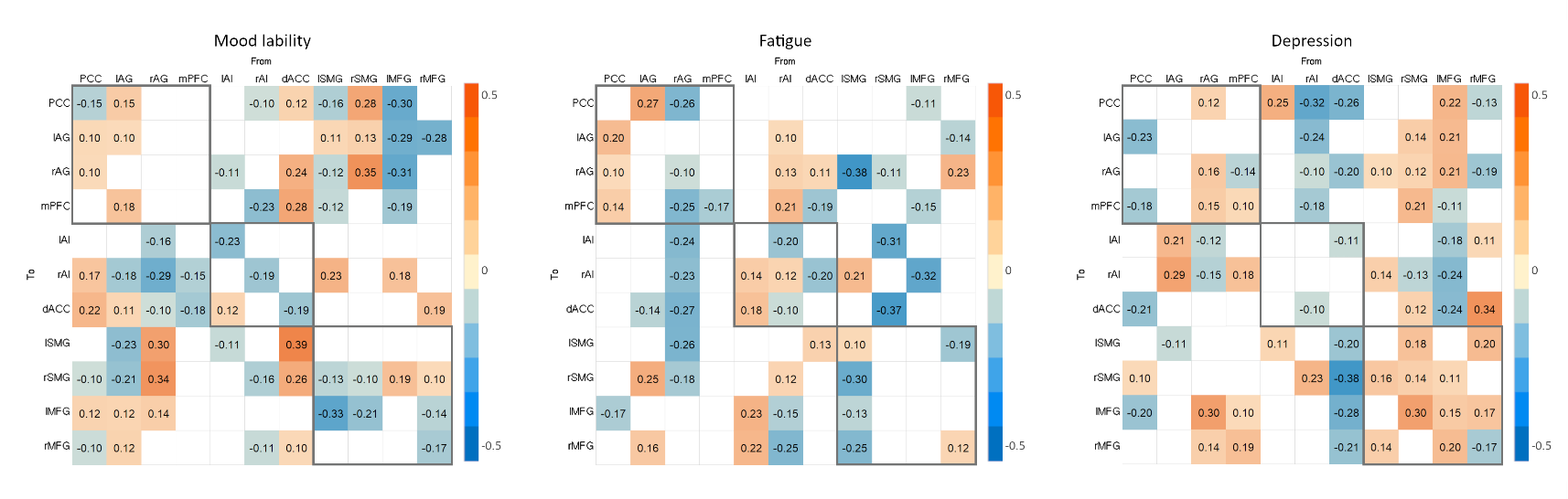

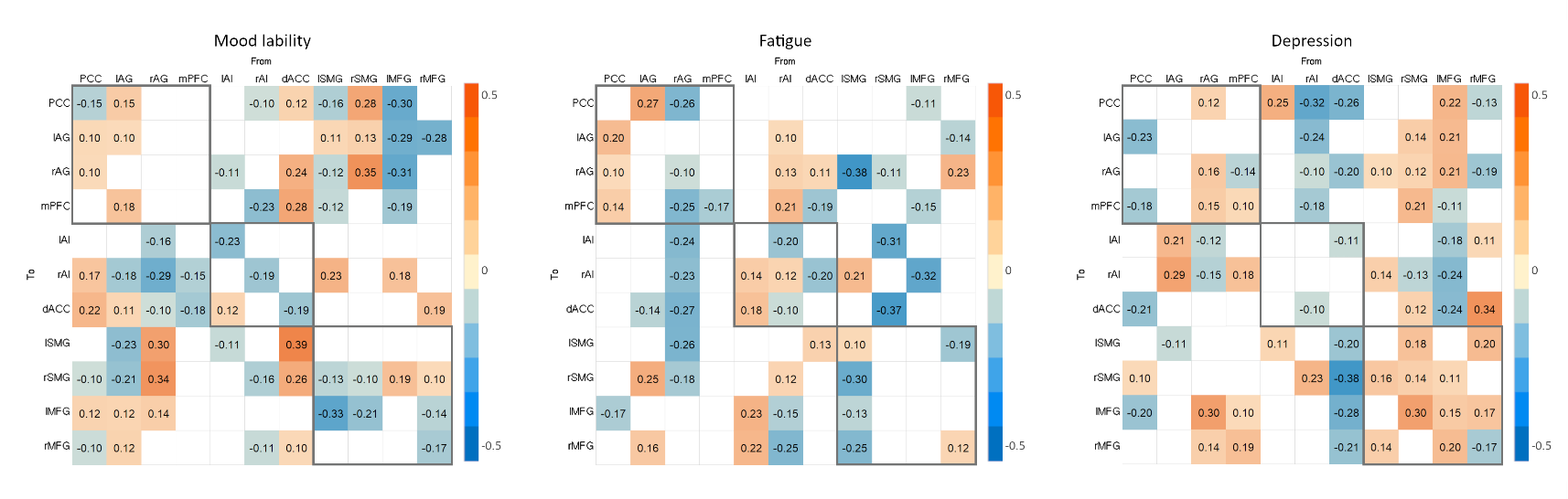

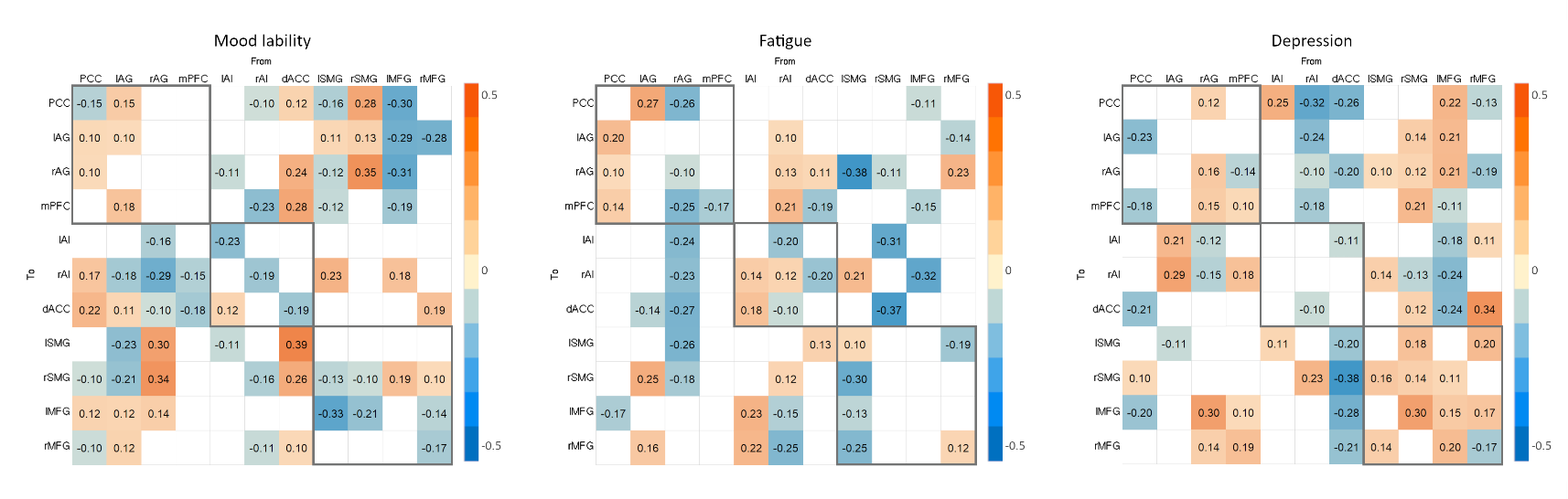

**Fig. S1. Estimated parameters for mood related symptoms.** These connections surpassed a posterior probability of 95% and an estimated value (Ep) of 0.10. The exact Ep is indicated in each cell, warm colours indicating positive parameter estimates and cold colours negative. The columns are the outgoing connections, and the rows are the incoming connections, ordered as: PCC, lAG, rAG, mPFC, lAI, rAI, ACC, lSMG, rSMG, lMFG, and rMFG.
